# COVID-19 lockdowns may reduce resistance genes diversity in the human microbiome and the need for antibiotics

**DOI:** 10.1101/2021.03.08.21253164

**Authors:** João S. Rebelo, Célia P. F. Domingues, Francisco Dionisio, Manuel C. Gomes, Ana Botelho, Teresa Nogueira

## Abstract

Recently, much attention has been paid to the COVID-19 pandemic, yet bacterial resistance to antibiotics remains a serious and unsolved public health problem, which kills thousands of people annually, being an insidious and silent pandemic. In this study, we explored the idea of confinement and the tightening of the hygiene measures to contain the spreading of coronavirus, to simulate the effect that it has on lowering the spreading of pathogenic bacteria in a human network, and on the need to use antibiotics. For that, we used computational biology to generate simulations.

## 3. Introduction

Antibiotic resistance is today one of the biggest threats in human and public health. It is a hidden pandemic! Antimicrobial-resistant pathogens already cause 25 000 deaths per year in Europe, 700 000 worldwide, and it is estimated that it could increase to 10 million by 2050, in the most alarming scenario if no action is taken (1). This also implies higher treatment costs due to the prolonged recovery time in biosafety facilities, but also the need to resort to more expensive alternative antibiotics (2).

Antibiotic misuse and over-prescription have long been documented as one of the most common errors in human health and one of the main factors that drive development of antibiotic resistance in hospitals. Misuse of antibiotics in humans can include unnecessary prescription, or treatments that are not streamlined when microbiological culture data become available (3,4).

Hygienic measures, such as hand washing, have long been recommended to break the transmission of pathogens, especially in hospitals and health care facilities. Recently, with the emergence of the new pandemic of COVID-19, new and stricter recommendations have been made, such as avoiding interpersonal contacts, disinfecting hands when touching physical surfaces in anthropogenic environments and following strict rules of respiratory etiquette, quarantine and confinement. It is therefore expected that, under such drastic hygienic conditions and interruption of the transmission of pathogenic microorganisms between people, antibiotic resistance in human microbiomes will also be reduced.

Human metagenomes with higher diversity of virulence genes tend to be precisely those with higher diversity of antibiotic-resistance genes (5) possibly facilitating the emergence of superbugs. However, the diversity of both gene types is dynamic over time, changing according to antibiotics usage and to microbial transmission between people (6). Bacterial transmission occurs between people that establish a network of contacts among them. What would happen to the diversity of antibiotic resistance genes if most contacts between people were abolished?

The aim of this study is to simulate the effect of human contacts reduction (person-to-person transmission in a network of contacts) on the accumulation of antibiotic resistance genes, due to the sharing of bacteria belonging to the human microbiota. In the same way, we intended to find out if it is to be expected that the decrease in human contacts could, by itself, explain a decrease in antibiotic intake in humans.

This study is based on data from a densely populated region of Portugal, the Lisbon and Tagus Valley region. Portugal was one of the European countries that most quickly managed to control its first wave of the covid-19 pandemic. Learning from the experience of other countries such as Spain or Italy, where SARS-CoV-2 caused many human losses, the Portuguese population massively followed the measures of isolation and social distancing enacted by their government to combat the pandemic.

The Lisbon and Tagus Valley region comprises a metropolitan area with about 2.855.000 persons over 3.000 Km^2^ (7) and the surrounding suburbs, with high daily traffic to and from Lisbon and with an overloaded transport network. For the development of this study, we used real data on the change in population mobility in this geographical region, immediately after the peak of the first wave, on the 1st of April, and extending until September the 30th, 2020.

## 4. Section heading(s)

### 4.1 Methods

The model used in this article is based on our previously developed model (6).

#### Data

In this work we used mobility data from Google (8) This data is made available to provide statistics on what has changed in response to policies aimed at combating COVID-19. The data are divided by region and discriminated into the following categories: grocery and pharmacy, parks, transit stations, workplaces and residential. For this study we used data from Portugal, namely from the Lisbon and Tagus Valley region, with approximately 3658623 individuals. Data were collected to correspond to the time period between April 1 and September 30, 2020 which includes a full lockdown period, between 1st April and 4th May, followed by a lighter one.

#### Building the human network

We simulated a network where each node represents a person or, more precisely, a person’s metagenome. The edges represent possible transmission avenues of microorganisms.

We built the social contact network following the Watts and Strogatz method (9) In the beginning of each simulation, we construct a regular network, in which each individual is connected to the n nearest nodes. Subsequently, we allow each connection to change with a probability (*p*) of 0.5. If it changes, the node will be connected to another node chosen at random. In this way, we obtain a small-world network, that remains unchanged in all cycles

#### Number of contacts

The distribution of the number of contacts per individual per day is not known in Portugal. We therefore used the data from the Polymod project, based on diaries of individual daily contacts in eight European countries (10). In this project, participants were chosen to register all the individuals with whom they had contact during a day. This provided an idea of the average number of daily contacts for each country analysed in the project.

In order to estimate the average number of daily contacts in Portugal, we assumed that mobility is correlated with the number of contacts. By comparing the mobility data of google from the different countries that participated in this project with the mobility data from Portugal, we found that the country’s most similar to Portugal are Belgium and Poland. In these countries, there are an average number of 12 and 16 daily contacts, respectively. Following this line of reasoning, in our simulations, we define that the average number of connections - which may or may not become a contact in a given day - between two individuals in the network (*n*) is 14.

#### Contacts per cycle

As previously mentioned, we initially established a small-world contact network. From the beginning of the simulation, each individual has an assigned set of different individuals to whom he or she will be connected during all the cycles. However, in each cycle, not all contacts take place as there may be a reduction in mobility (in percentage). This reduction is reflected in an equal percentage decrease in the number of contacts that each individual has. For example, if with 100% mobility (base line) an individual is connected to 10 other individuals, with 60% mobility in one cycle, the individual will be connected in that cycle to only 6 other individuals. We assume that the connections occurring preferentially are the ones with the closest individuals in the network (representing, for example, the household). For instance, if the connections of individual 1 defined initially are with individuals 2, 4, 5, 9, 18, 22, 27, 35, 48, 87 (ten connections), if there is 60% mobility (a reduction of 4 connections), the connections that do not occur will be with individuals 27, 35, 48 and 87.

In the simulations, we considered three mobility scenarios: (i) using the real mobility data from Google from the period between April 1st and September 30th; (ii) with a fixed mobility of 60%; and (iii) with a fixed mobility of 40%.

#### The metagenome, pathogenic bacteria, and antibiotic administration

The model considers the transmission of bacterial pathogens (capable of causing infections), as well as antibiotic resistance genes, between people. These genes are present in the metagenome. We focused on the presence or absence of genes encoding different functions, irrespectively of its copy-number in the metagenome. In the simulations, each gene represents a gene family (with similar functions). We divided resistance genes into groups, each group having the same number of families. Each group represents genes associated with resistance to an antibiotic. Of note, we did not consider resistance to multiple drugs in our simulations. Therefore, there will be as many groups as there are antibiotics accounted for in the simulations. We define the diversity of a specific gene kind as the number of genes of that type present in a human metagenome.

To simulate the migration of bacteria from individuals outside the network or the ccontamination from sources such as food or contaminated water, we inserted some different bacterial pathogenic species into random individuals per cycle. In this model, the only difference that we consider between species is the antibiotic to which they are susceptible, as explained below.

Individuals infected by pathogenic bacteria feel sick and take an antibiotic. The antibiotic administered is specific for the bacteria that caused the infection disease. The antibiotic selects cells carrying resistance genes by eliminating the remaining susceptible bacteria. We assume that all families of resistance genes are present in all metagenomes, but in two different possible states: in some metagenomes, they are present in low copy number, so they are not likely to be transmissible to other individuals in the network; in other metagenomes, the copy number of resistance genes is high due to the selective pressure of antibiotics to which they were previously submitted. In the latter case, resistance genes are likely to be transmissible from person to person.

Moreover, upon antibiotic consumption, the following events can occur: (i) elimination of susceptible pathogenic bacterial species; (ii) selection of resistance genes belonging to the group of resistance to that antibiotic (which means their copy number gets so high that they become transferable); (iii) loss of resistance genes associated with other antibiotics with a given probability (becoming non-transferable but still present in minute copy number).

Several processes lead to gene loss. Genes are lost because of the selective pressure by antibiotics and because we assume that resistance determinants impose a fitness cost (in the absence of antibiotics). To include this cost in the simulations, we consider that each metagenome may lose specific resistance genes according to a “loss rate” (with this process, these genes become non-transferable).

#### Algorithm of the program

Each simulation is composed of several cycles, where each one corresponds to a day. In each cycle, we considered all procedures described in the pseudocode (Figure 1). We used as default the parameterized values of our model (6). The main steps of the program in each cycle are:

**Figure 1.**
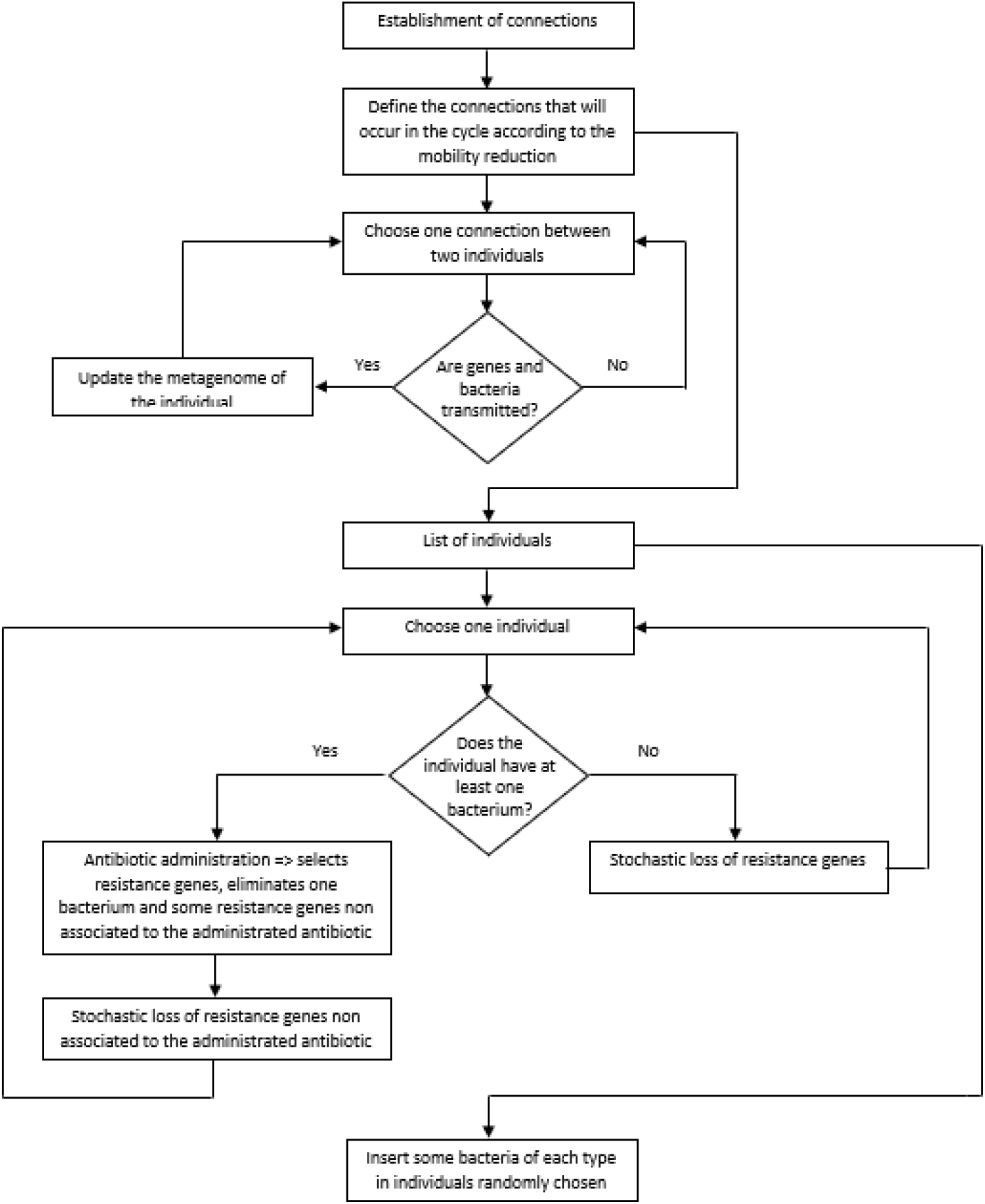
Flowchart of the algorithm of the program.

i. Choose the connections that will occur in that cycle according to the percentage of mobility in that cycle (as explained above).
ii. Transfer of pathogenic bacteria and resistance genes between people (i.e., between linked nodes in this cycle).
iii. Look for people infected by at least one pathogenic bacterial species. These people take antibiotics (chosen according to the pathogen). The antibiotic eliminates the pathogenic species and selects the resistance genes associated with the antibiotic used. The antibiotic also eliminates resistance genes unrelated to the administered antibiotic. Finally, the metagenome loses a few more resistance genes not associated with the antibiotic. The cause of this loss is the fitness cost of resistance genes.
iv. The metagenomes of people that did not take an antibiotic in this cycle lose resistance genes. This loss is a consequence of the fitness cost imposed by resistance genes on their hosts.
v. Insert five bacterial pathogenic species in individuals randomly chosen from the population.

#### Statistical analysis

In each simulation we counted the number of resistance genes of each individual. From these data, we constructed a graph that represents how many individuals (vertical axis) have each amount of resistance genes (horizontal axis) at the end of the 180 cycles. A Kruskal-Wallis test was used to compare the effect of different confinement regimes, followed by a Dunn post hoc analysis. We performed both tests with R – version 3.5.1 (11) and used the FSA package (12) to perform the Dunn’s test.

### 4.2 Results and Discussion

#### Lockdown breaks many human contacts on the social network

In Portugal the first pandemic wave COVID-19 occurred between March and August 2020. From the 16th of March, the Portuguese authorities decreed a state of emergency and a global lockdown. The Lisbon and Tagus Valley area in Portugal is a predominantly urban area that showed greater variations in circulation, and therefore in human networks, before and during lockdown due to the COVID-19 pandemic.

According to Google data (8), the mobility was very low in the beginning of April and increased mainly since May with many oscillations (Fig.2). The lower mobility registered through Google data, gives us an indication of the decrease in the number of contacts between individuals. It is, however, only an estimate of the human social contacts, since it contemplates only individuals who move with connected mobile data on and does not distinguish between individuals who move and contact other people from another household, from those who move lonely complying the social distancing rules.

During lockdown, intimate human connections should remain preserved at home. However, we expect a reduction in the transmission of bacteria and pathogens between individuals in the community and, therefore, less bacterial enrichment of human microbiomes. As a consequence, we can predict a reduced flow of antibiotic-resistant bacterial genes between people’s microbiomes during confinement (13).

Likewise, mobility, as a measure of the number of times an individual leaves home, is not the only factor that influences the exchange of microorganisms in a social network of contacts. The use of a mask and other behaviours like social distancing, also reduce the spread of pathogens, particularly in the case of airborne disease agents.

#### Confinement leads to lower diversity of antibiotic resistance genes in human microbiomes

In order to test the effect of the confinement in reducing the transmission of antibiotic resistance genes (ARGs) in the human microbiomes of the population, we simulated four different scenarios: *i*) no confinement, hence no connections eliminated; ii) using the real mobility data from Google that comprises all the period of time of the first pandemic wave in Portugal assuming that a certain decrease of mobility corresponds to the same decrease of the number of connections; and assuming constant reductions of connection between individuals of *iii*) 40% and *iv*) 60%.

According to Google data, mobility fluctuated over time, ending with a mobility of the individuals nearly 90%, consistent with the end of the first pandemic COVID-19 wave and summer holidays (Fig. 2). Also, according to Google, mobility was, in average, 75% of the maximum during this period of time.

**Figure 2.**
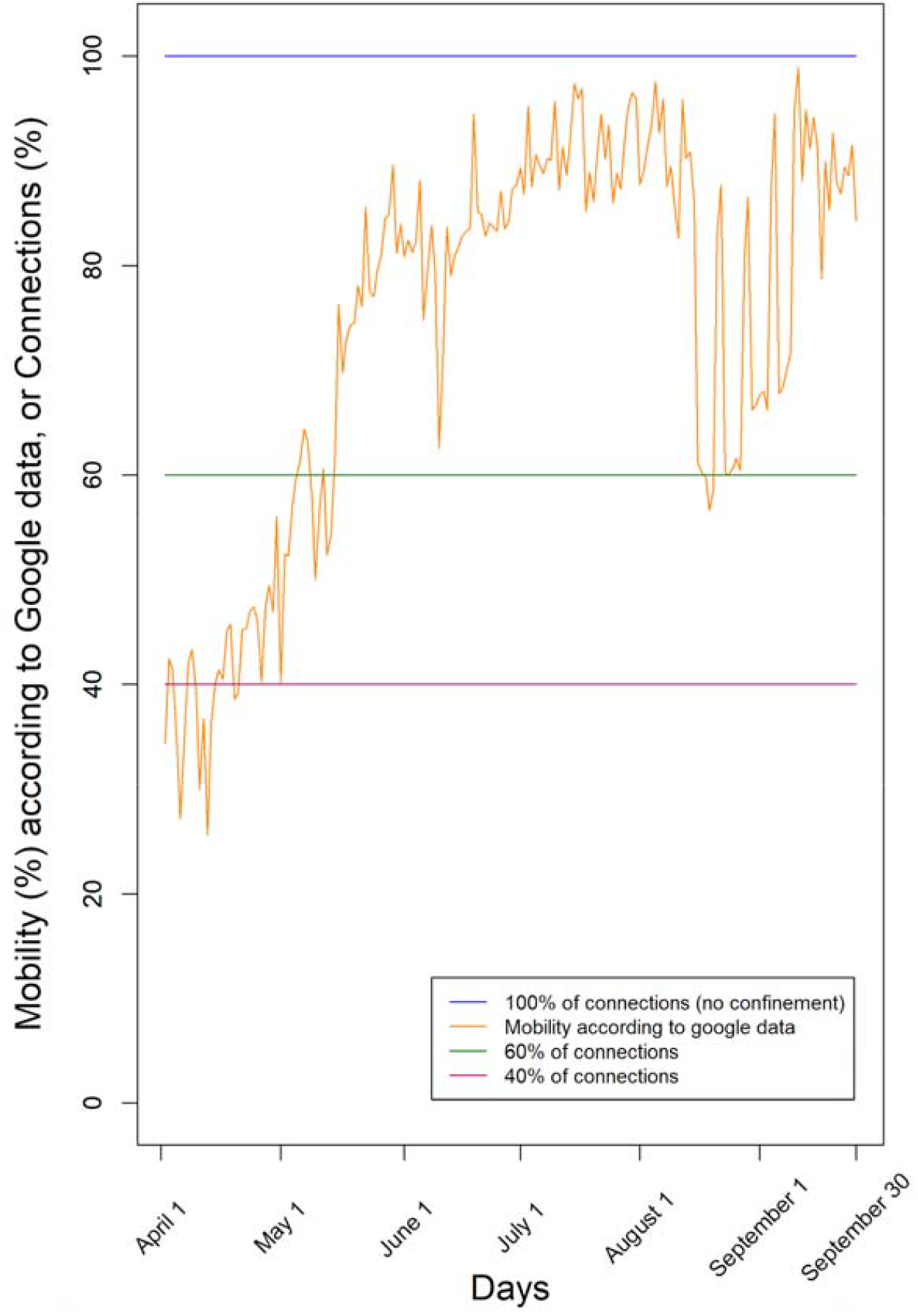
Levels of confinement used in the simulations. We used either a fixed number of connections (100%, 60%, or 40%), or mobility according to Google data from the region of Lisbon and Tagus Valley in Portugal in the period April 1st - September 30th.

Our simulations of a 180-days period generated the distributions of the diversity of resistance genes in the population shown in Fig. 3. The modes of the four distributions are: 90 gene types for the case of no restrictions to connections, 83 assuming that google data corresponds to connections relative to the normal situation, 75 if the population only has 60% of contacts, and 59 if the population only has 40% of contacts.

**Figure 3.**
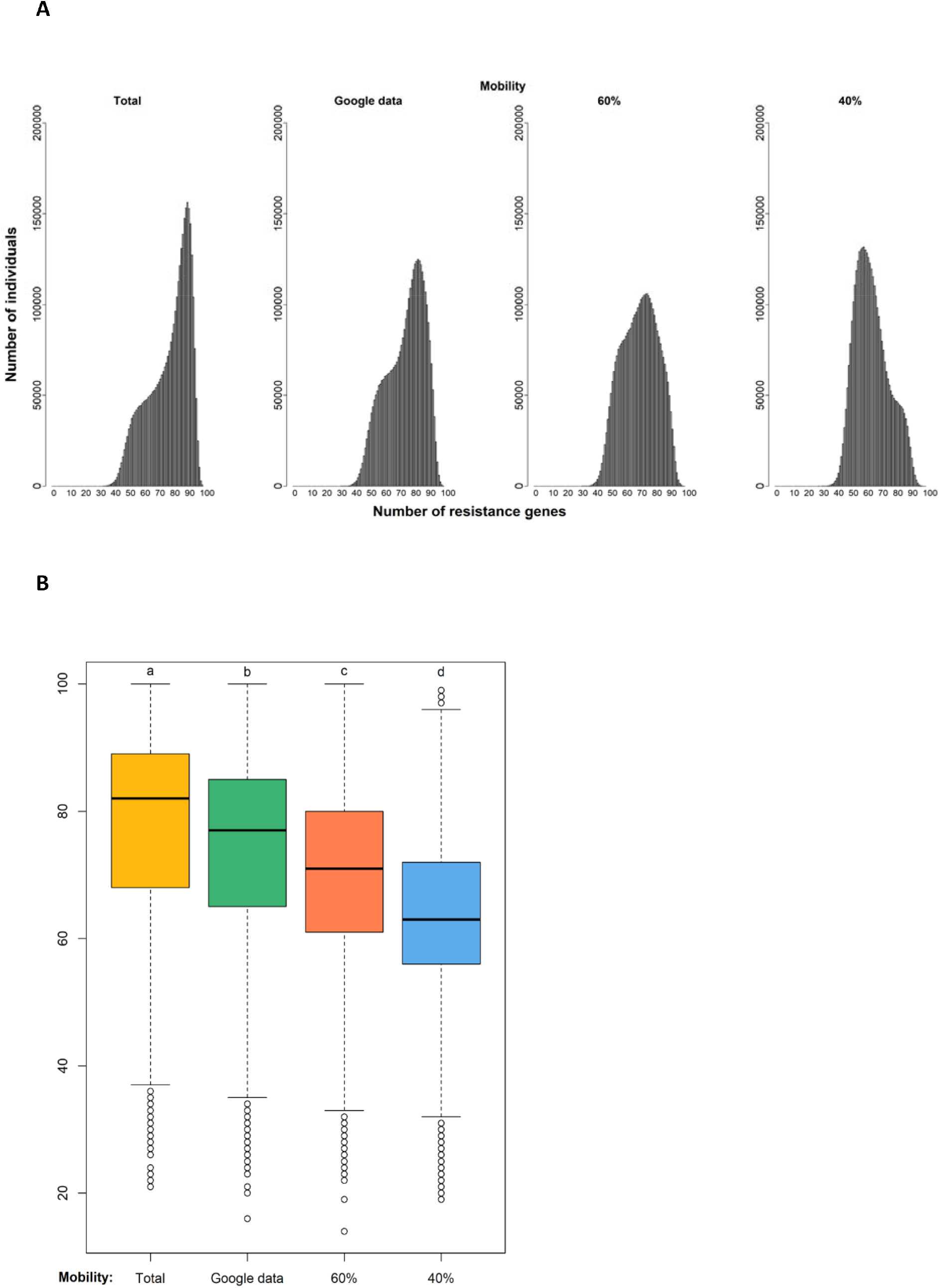
Diversity of resistance genes in four different scenarios of confinement after a 180-day period. Simulation (i) without confinement (100% of connections); (ii) using the real mobility data from Google assuming that a certain reduction of mobility corresponds to a similar reduction of connections; and with a fixed proportion of connections of (iii) 60% and (iv) 40%. A: Distribution of the predicted number of antibiotic resistance genes types among individuals at the end of simulation. B: Each boxplot represents the distribution of values: the bottom and top of the box are the first and third quartiles, the horizontal line is the median, the vertical lines are the 1.5 interquartile range. Black circles represent outliers. After a Kruskal-Wallis analysis (see main text), post hoc comparisons using the Dunn test indicated that the effect of confinement is significantly different in the four confinement types as indicated by the four different letters a,b, c and d (p-value<.000).

A Kruskal-Wallis test provided evidence of a difference (p < .000) between the mean ranks of at least one pair of groups. Dunn’s pairwise tests were carried out for the six pairs of groups. There was very strong evidence (p < .000, adjusted using the Bonferroni correction) of a difference between the four confinement regimes. The proportion of variability in the ranked dependent variable accounted for by the confinement variable was η^2^ = .148, indicating a large effect of confinement on the diversity of antibiotic resistance genes.

#### Less contact between humans reduces the need for antibiotic use

According to our algorithm, the use of antibiotics is triggered by exposure to bacterial pathogens during social interactions. When running the simulation, we assume that when an individual is more exposed to bacteria from other individuals, as part of a larger network of contacts, there is an increased likelihood of being infected by a pathogen and taking antibiotics. Thus, we counted the number of antibiotic administrations that occurred during the simulation. Compared with the unconfined condition, the number of antibiotic administrations fell 7, 11 and 16% in the other three conditions: google data (assuming that mobility loss corresponds to the same connectivity loss), 60% of connections, and 40% of connections. Therefore, the daily use of antibiotics seems to follow a similar trend pattern of confinement. This trend may explain, at least partially, why, according to data from Infarmed (Portuguese drug agency), the amount of antibiotics used in Portugal in the period between January and September 2020 was lower than the corresponding months of 2019 (11). In community pharmacies, the dispensing of these drugs decreased by 20%. However, the reduction in prescription and use of antibiotics may reflect a lower need for these drugs, but also a decrease in the number of medical appointments and access to primary health care due to the pandemic.

### 4.3 Conclusions

In the work, we have shown that a reduction of contacts between people leads to a general lower diversity of antibiotic-resistance genes among individuals’ metagenomes.

In our simulations, we assume that interpersonal contact enriches the human microbiome with genes, namely those that confer resistance to antibiotics, but also leverages the risk acquisition of pathogens which, in turn, triggers the taking of antibiotics. Thus, it seems easy to understand that the reduction in mobility that occurred during a period of confinement, in our simulations, led to a decrease in bacterial infections in humans, and concomitantly to a decrease in the prescription of antibiotics, as well as to a decrease in the accumulation of antibiotic resistance genes in the microbiome. This result is concordant with the 20% drop in the prescription of antibiotics in the first three quarters of 2020 reported by the Portuguese National Health Authorities (14). This conclusion reinforces the general recommendations to improve hygiene in disease control and antibiotic resistance in human populations.

For the parameterization of the model developed in this work, we have used data from a mixed origin: close-to-real data of the population of a geographical area, data that was estimated from the bibliography, for example the network of contacts, or theoretical data, such as the probability of an individual taking antibiotics. However, during the development of this work, we only varied one of the parameters, namely the changes in the human contact network as an indicator of the spread of pathogens among different individuals. Thus, we consider that the conclusion that the breakdown of human contacts leads to decrease in the enrichment of the human microbiome in antibiotic genes is significant and is also supported by reality.

## Data Availability

There are no new data generated

## 5. Author statements

### 5.1 Authors and contributors

Conceptualization: JR, CD, FD and TN; Formal Analysis: JR, CD, FD, MCG and TN; Investigation and Methodology: JR, CD, FD and TN; Project administration and Supervision: FD and TN; Software: JR and CD; Visualization: JR, CD, FD and TN; Writing – original draft: JR, CD, FD and TN; Writing – review & editing: JR, CD, MCG, FD, AB and TN

## 5.4 Acknowledgements

We thank Octávio Serra for technical help and Fernanda Simões for granting access to the servers. T.N. was supported by contract ALG-01-0145-FEDER-028824. Fundação para a Ciência e a Tecnologia (FCT), I.P., supports cE3c through contract UIDP/00329/2020

